# Risk of cardiovascular disease in patients with classical Hodgkin lymphoma: a Danish nationwide register-based cohort study

**DOI:** 10.1101/2024.09.30.24314272

**Authors:** Sissel J. Godtfredsen, Harman Yonis, Joachim Baech, Nour R. Al-Hussainy, Signe Riddersholm, Lars Kober, Morten Schou, Jacob Haaber Christensen, Martin Hutchings, Rasmus Bo Dahl-Sørensen, Peter Kamper, Caroline E. Dietrich, Mikkel Porsborg Andersen, Christian Torp-Pedersen, Peter Sogaard, Tarec Christoffer El-Galaly, Kristian H. Kragholm

**Affiliations:** Department of Cardiology, Aalborg University Hospital, Aalborg, Denmark; Department of Cardiology, Nordsjaellands Hospital, Hilleroed, Denmark; Department of Public Health, University of Copenhagen, Denmark; Department of Hematology, Clinical Cancer Research Center, Aalborg University Hospital, Aalborg, Denmark; Department of Cardiology, Copenhagen University Hospital – Rigshospitalet, Copenhagen, Denmark; Department of Cardiology, Copenhagen University Hospital – Herlev and Gentofte Hospital, Hellerup, Denmark; Department of Hematology, Odense University Hospital, Odense, Denmark; Department of Hematology, Copenhagen University Hospital, Rigshospitalet, Copenhagen, Denmark; Department of Clincal Medicine, University of Copenhagen; Department of Hematology, Zealand University Hospital, Roskilde, Denmark; Department of Hematology, Aarhus University Hospital, Aarhus, Denmark; Department of Medicine Solna, Clinical Epidemiology Division, Karolinska Institutet, Stockholm, Sweden; The Prehospital Center, Region Zealand, Naestved, Denmark; Department of clinical medicine, Aalborg University, Aalborg, Denmark

## Abstract

**Aim:** To describe the risk of cardiovascular disease (CVD) in patients with classical Hodgkin lymphoma (cHL) undergoing contemporary treatment.

**Methods:** Including all patients with cHL ≥18 years at diagnosis treated with doxorubicin-containing chemotherapy between 2000-2022. Patients were matched with comparators in a 1:5 ratio on birth year, sex, and Charlson Comorbidity Index at time of matching (score of 0 or ≥1). A composite of CVDs including coronary artery disease, heart failure, valvular stenoses, restrictive pericarditis, arrythmias, or any procedures related to these was the primary outcome. Follow-up started on date of cHL diagnosis (i.e., the matching date). The cause-specific cumulative incidence and 95% confidence intervals (CIs) were computed with all-cause death and lymphoma relapse as competing events using the Aalen-Johansen estimator.

**Results:** A total of 1,905 patients with cHL and 9,525 comparators were included. Median age was 39 years (interquartile range, [IQR]: 27-56), 58% were males, most were treated with ABVD (83%), and the median cumulative doxorubicin dose was 250 mg/m^2^ (IQR: 200-300). Median follow-up was 10 years (IQR: 5.9-17.4). The cumulative incidences of CVD were 4.7% (95% CI: 3.6-5.7) for patients vs. 2.6% (95% CI: 2.3-2.9) for comparators at 5 years, 8.9% (95% CI: 7.2-10.5) vs. 5.5% (95% CI: 4.9-6.0) at 10 years, and 17.0% (95% CI: 14.1-19.9) vs. 8.2% (95% CI: 7.4-9.0) at 15 years.

**Conclusions:** CVD remains a substantial late effect after contemporary treatment for cHL, suggesting that awareness of cardiac symptoms and a low threshold for referral to diagnostic examination are still important measures during survivorship.

## Introduction

Survivorships issues are crucial in patients with classical Hodgkin lymphoma (cHL) due to cure proportions exceeding 80% and a young median age at diagnosis.^1,2^ In the past decades, several studies have reported significantly increased risk of late complications after treatment of cHL, with secondary malignancies and cardiovascular diseases (CVD) being particular concerns.^3–7^ Cardiovascular complications after cHL include an elevated risk of coronary artery disease, valvular diseases (particularly left sided), arrhythmias, heart failure, cardiomyopathies, pericardial disease, and autonomic dysfunction.^8–14^ The risk is substantial with a cumulative incidence of CVD as high as 50% over a 40-year period,^15^ with coronary artery disease, heart failure, and valvular disease being the most common cardiovascular complications with 20-year incidences of 10% and 6%.^9^ A Dutch study reported that the risk increase translated into an additional 62 cases of coronary artery disease per 10,000 persons per year compared to the general population.^14^ However, most studies on treatment-related cardiotoxicity have focused on patients treated between the 1960s and late 1990s. During that period, mantle field and other extended radiation fields covering the mediastinum/chest were frequently used.^8–12,15^ The radiation to the heart associated with these techniques has been speculated to be the single most important contributor to excess cardiovascular disease in cHL survivors treated in that era.^8,9,14,16^ During the past decades, advancements in radiotherapy techniques have limited heart radiation doses, along with a decreased use of radiotherapy.^17^ It is now primarily used in patients with limited stage disease and a minority of patients with advanced stage disease and F-18 fluorodeoxyglucose (FDG)-positive residual lymphomas.

While cardiovascular complications after modern cHL treatment are expected to decrease, patient remain at increased risk due to the widespread use of anthracycline-based chemotherapy regimens in the frontline setting.^18^ Historically, 550 mg/m^2^ of doxorubicin has been the commonly cited maximal tolerated dose avoiding a very high risk of cardiotoxicity.^19^ The maximum cumulative dose of anthracyclines used in first line treatment of cHL today is 300 mg/m^2^ corresponding to 6 cycles of doxorubicin-bleomycin-vinblastine-dacarbazine (ABVD) and therefore well below this threshold. Nevertheless, studies of non-Hodgkin lymphoma have shown substantial increase in risk of heart failure in patients receiving less than 300 mg/m^2^.^20^ The purpose of this population-based study was to investigate patterns of cardiotoxicity following modern treatment of cHL and to describe clinicopathologic features associated with higher risk of cardiotoxicity among patients with cHL.

## Methods

### Study design and setting

This was a nationwide cohort study with the following inclusion criteria: (1) cHL diagnosis between January 1^st^, 2000, and December 31^st^, 2022; (2) age ≥18 years at diagnosis; and (3) received at least one cycle of ABVD or bleomycin-etoposide-doxorubicin-cyclophosphamide-vincristine-procarbazine-prednisolone (BEACOPP) Patients with missing number of chemotherapy cycles were excluded.

Each patient was matched 1:5 to an individual from the Danish population free from cancer on the date of the patient’s diagnosis on year of birth, sex, and Charlson comorbidity index (CCI, 0 or ≥1, i.e., no comorbidities or one or more comorbidities at the time of matching) using risk set matching method. Patients and comparators with heart or cancer disease prior to the index date were excluded.

### Data sources

The Danish health care system is tax-based with free services for all residents.^21^ Data concerning healthcare use is collected by the Danish authorities in various registers. Linkage between registries is possible on an individual level through the civil personal registration (CPR) number assigned to all Danish residents at birth or immigration.^22^ Multiple registries were used in this study. The National Lymphoma Registry (LYFO) contains detailed information on all lymphoma patients in Denmark since 2000. LYFO provided information on treatment (type and courses of chemotherapy, detailed information on radiotherapy if applicable), clinicopathological features at the time of diagnosis, and treatment outcomes (including relapse and death).^23^ The Danish National Patient Registry provided data on hospital admissions, discharge diagnoses, and procedures using the International Classification of Diseases (ICD) system and the Nordic Medico Statistical Committee (NOMESCO) classification.^24^ The Danish National Prescription Registry provided data on all filled prescriptions by Anatomical Therapeutic Chemical (ATC) codes.^25^ The Danish Registry of Causes of Death provided information on deceased individuals.^26^ The Danish Population Education Registry provided information on highest achieved level of education at the time of matching.^27^ International Standard Classification of Education (ISCED) was converted to a score of 1-4 where 1 indicates ISCED levels 0-2, 2 indicates ISCED level 3, 3 ISCED levels 5-6, and category 4 equals ISCED levels 7-8, as done previously.^28^

### Outcomes

The primary outcome was a composite endpoint of several cardiovascular diagnoses and procedures: coronary artery disease, acute coronary syndrome, heart failure, aortic-, mitral- or tricuspid stenosis, cardiomyopathy, restrictive pericarditis, or atrial or ventricular arrythmias (from this point referred to as cardiovascular disease [CVD], ICD-10 diagnosis codes and NOMESCO procedure codes are summarized in *Supplementary material S1*). Secondary outcomes included the individual diagnoses and procedures listed above and death from cardiovascular causes.

### Statistical analysis

For continuous variables, medians and interquartile range (IQR) were calculated, and for categorical variables frequencies and percentages. Cell counts ≤3 were reported as not applicable (NA) throughout.

Cause-specific cumulative incidences of CVD were estimated with deaths from any cause (except cardiovascular) and lymphoma relapse as competing events using the Aalen-Johansen estimator. Follow-up was from diagnosis (matching) date until date of first CVD event, emigration, death, or end of study (December 31^st^, 2022), whichever occurred first.

Patients with cHL were divided into subgroups according to treatment regimen: ABVD, BEACOPP, mixed ABVD/BEACOPP, and other treatments. The mixed ABVD/BEACOPP group was defined by at least 2 cycles of both ABVD and BEACOPP. The “other” group was comprised of patients receiving 1 course of ABVD or BEACOPP and more than 1 course of a different treatment regimen. As LYFO does not capture dosing information, total doxorubicin dose was estimated by assuming 50 mg/m^2^ per cycle of ABVD or cyclophosphamide, doxorubicin, (etoposide), vincristine, and prednisolone (CHO[E]P) and 35 mg/m^2^ per cycle of BEACOPP.

As an exploratory analysis, multivariable Cox proportional hazards models were fitted, yielding hazard ratios (HRs) with 95% confidence intervals (CIs), to identify clinicopathological features associated with cardiovascular disease among patients with cHL and comparators. The preselected risk factors included age, sex, hypertension, diabetes, disease stage at diagnosis, bulky disease, Eastern Cooperative Oncology Group (ECOG) performance status, hypercholesterolemia, cumulative doxorubicin doses, and radiotherapy. All factors were adjusted to age at cHL diagnosis, sex, and CCI (0 or ≥1). To further explore the effect of cumulative doxorubicin dose, a Cox proportional hazards model was fitted with a cubic spline transformation of cumulative doxorubicin dose (5 degrees of freedom) adjusted to sex and age at cHL diagnosis.

SAS version 9.4 (Cary, NC, USA) and R version 4.2.1 (R Development Core Team) were used for data management and statistical analyses, respectively.

### Ethics

In Denmark, register-based studies performed for the sole purposes of statistics and scientific research do not require ethical approval nor informed consent. The study was registered in the Capital Region of Denmark (P-2023-320) in compliance with the Danish Data Protection Act and the General Data Protection Regulation.

## Results

### Patients and characteristics

Between 2000 through 2022, 2,617 patients were diagnosed with cHL. Of those, a total of 712 were excluded due to: previous cancer (n=85), previous heart disease (n=177), no ABVD or BEACOPP treatment (n=343), missing number of chemotherapy cycles (n=101), and/or no Danish CPR number (n=6) (*Figure 1* consort diagram) leaving a final cHL population of 1,905 patients (*Table 1*).

**Figure 1:**
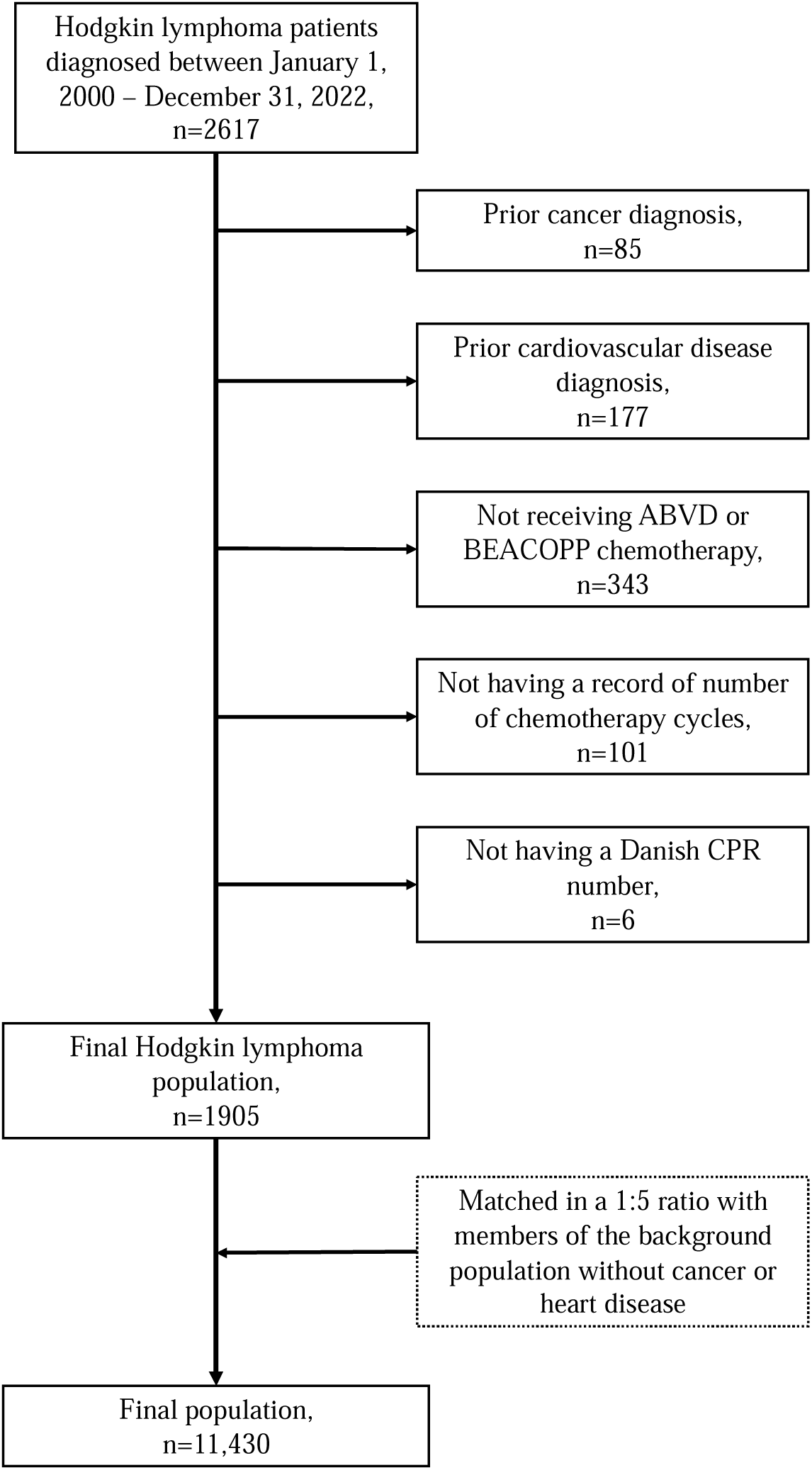
Flowchart summarizing the inclusion, exclusion, and final study population size. Abbreviations: CPR: civil personal registration; ABVD: doxorubicin-bleomycin-vinblastine-dacarbazine; BEACOPP: bleomycin-etoposide-doxorubicin-cyclophosphamide-vincristine-procarbazine-predinisolone

**Table 1:**
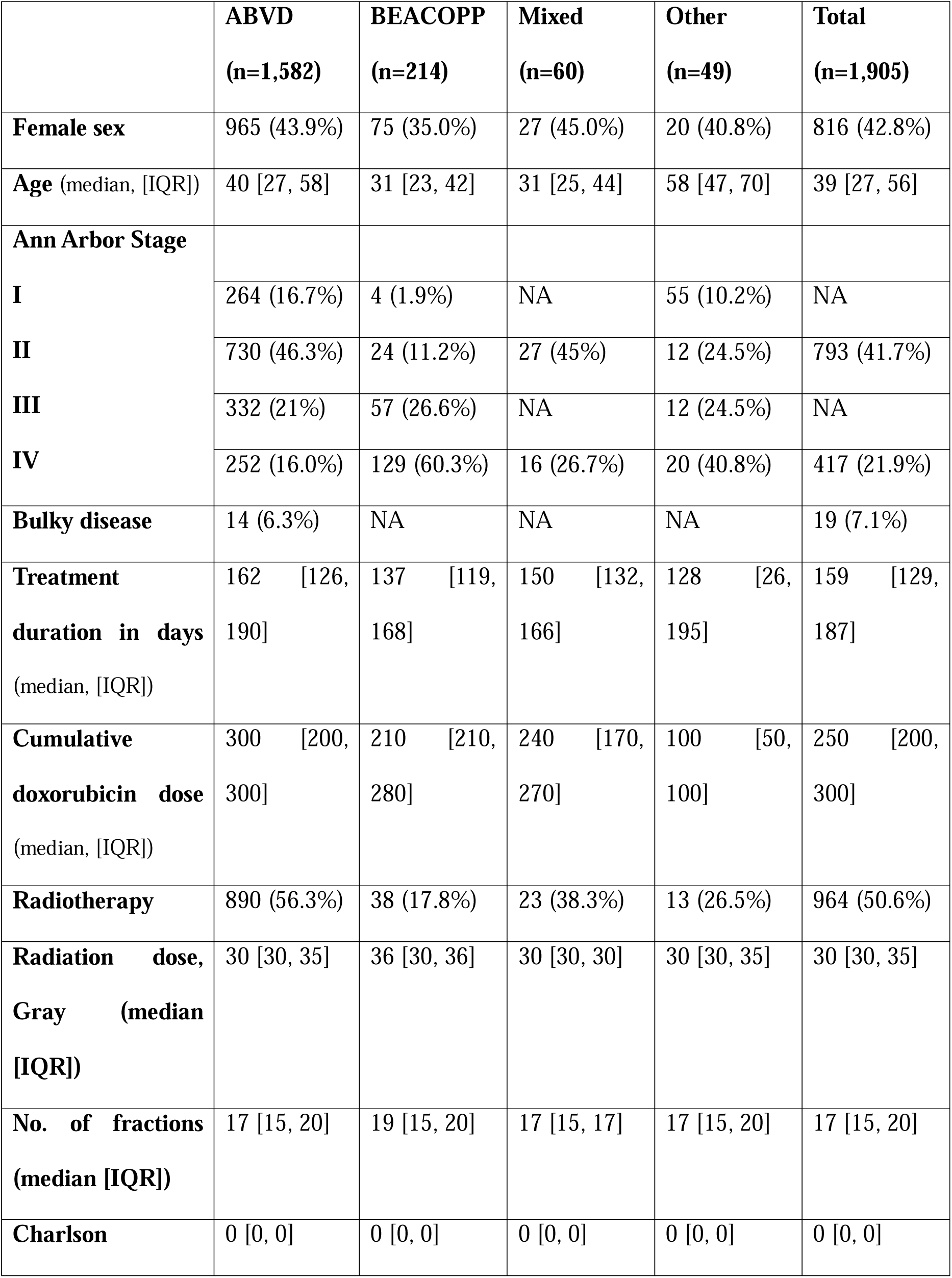
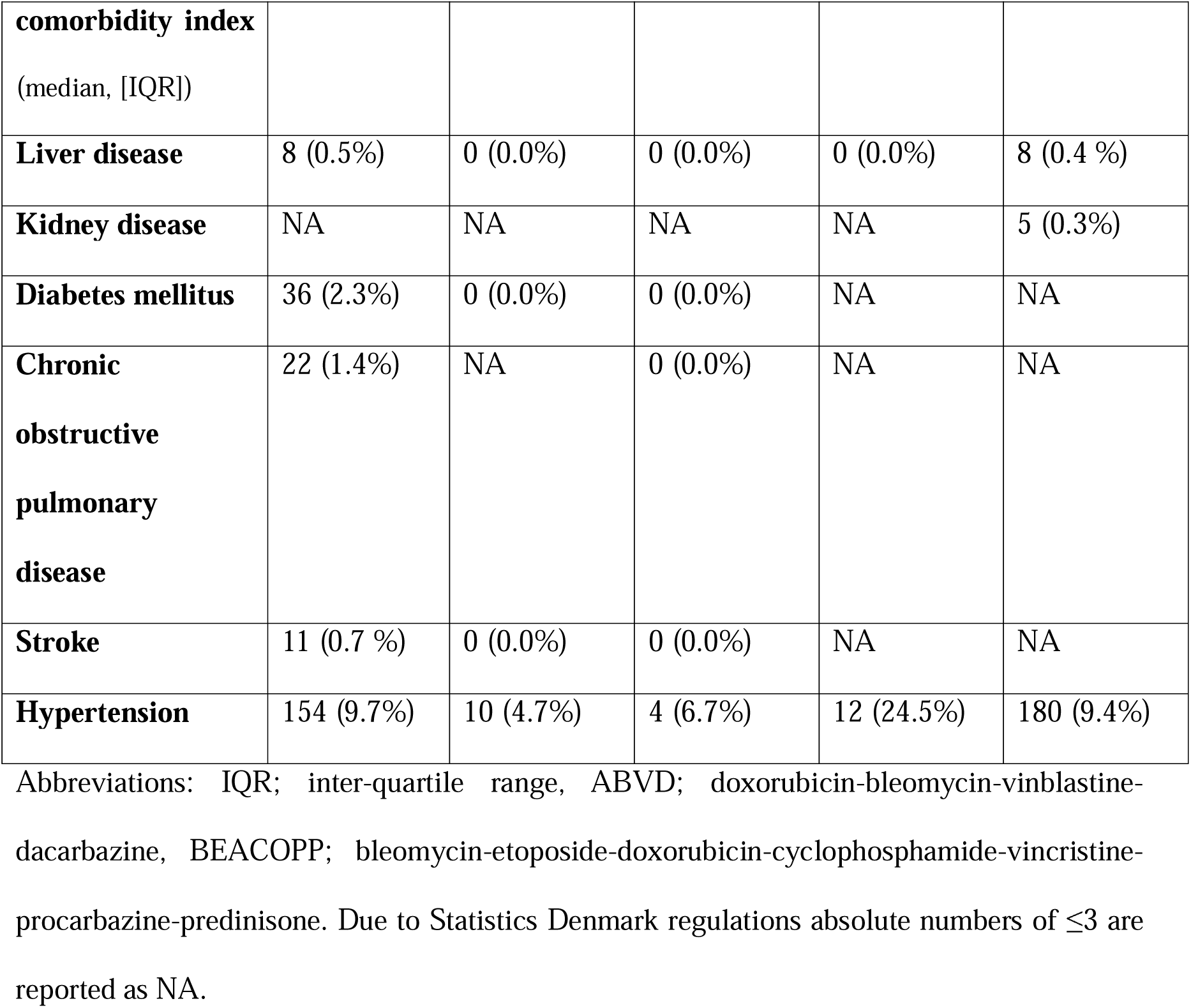
Baseline characteristics of the Hodgkin lymphoma patients stratified by treatment group.

Median age at cHL diagnosis was 39 (IQR: 27-56) and 57.2% were males. The majority of patients were treated with ABVD (83%), 11.2% with BEACOPP, 3.2% received both ABVD and BEACOPP (mixed), and 2.6% received only one cycle of ABVD or BEACOPP in combination with other treatments (other). The median cumulative doxorubicin dose was 250 mg/m^2^ (IQR: 200-300). Median doxorubicin dose for patients treated with ABVD was higher than for patients treated with BEACOPP (300 [IQR: 200-300] vs. 210 [IQR: 210-280], respectively). Approximately half of the all patients received consolidating radiotherapy (ABVD 56%, BEACOPP 18%, mixed 28%, and other 27%) with a median total dose of 30 Gray ([Gy], IQR: 30-35) and 28% of all radiotherapy-treated patients received 35 Gy or more. The most common radiation regimes were involved field and involved node (97%). During the follow-up period, 242 patients had lymphoma relapse.

Together with the 9,525 matched comparators the final study population comprised 11,430 individuals. Baseline characteristics of patients with cHL and matched comparators are presented in *Table 2*.

**Table 2:**
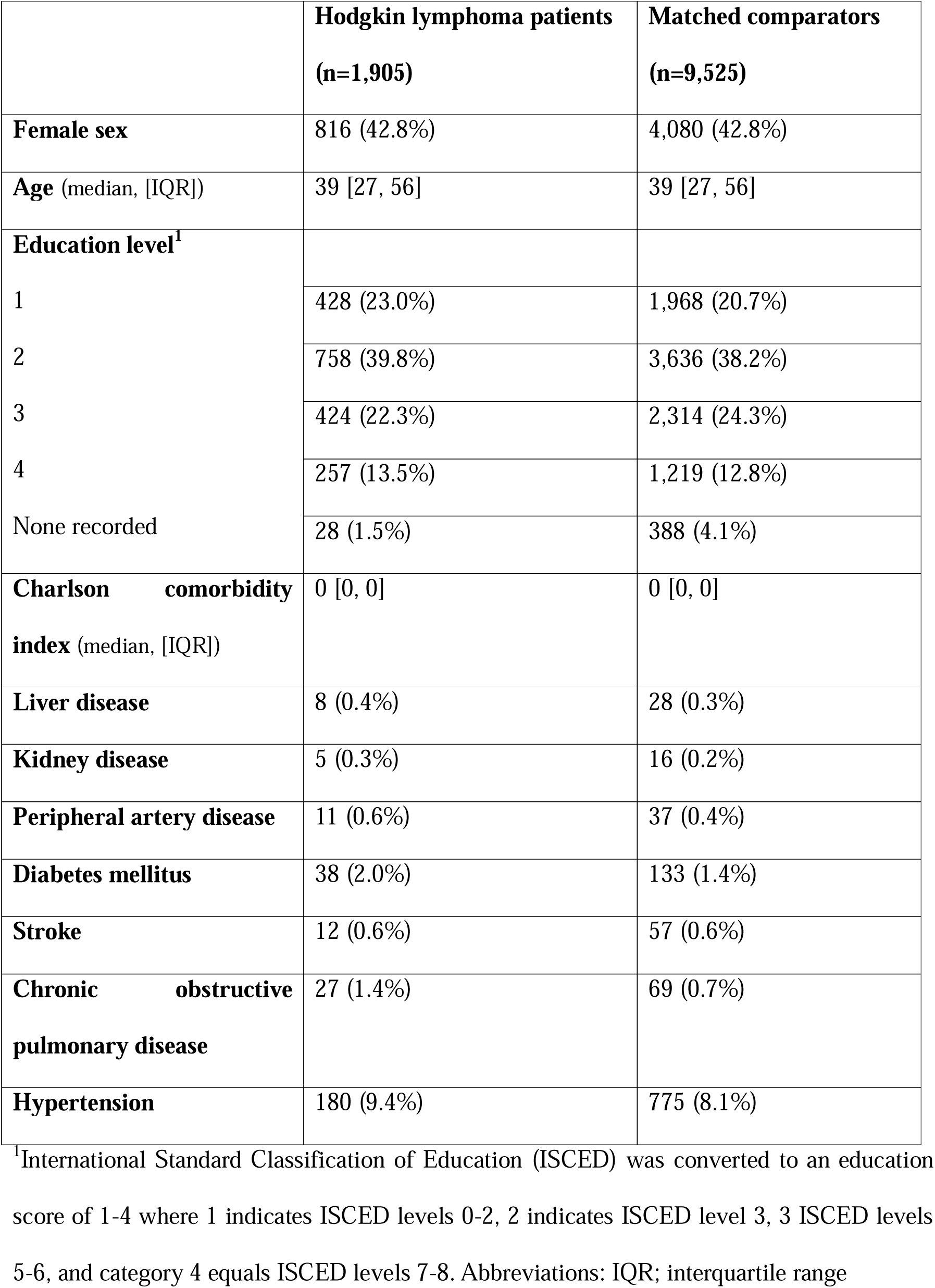
Baseline characteristics of the final population stratified by Hodgkin lymphoma patients and matched comparators.

|  | **Hodgkin lymphoma patients**<br>**(n=1,905)** | **Matched comparators**<br>**(n=9,525)** |
| --- | --- | --- |
| **Female sex** | 816 (42.8%) | 4,080 (42.8%) |
| **Age** (median, [IQR]) | 39 [27, 56] | 39 [27, 56] |
| **Education level<sup>1</sup>** |  |  |
| 1 | 428 (23.0%) | 1,968 (20.7%) |
| 2 | 758 (39.8%) | 3,636 (38.2%) |
| 3 | 424 (22.3%) | 2,314 (24.3%) |
| 4 | 257 (13.5%) | 1,219 (12.8%) |
| None recorded | 28 (1.5%) | 388 (4.1%) |
| **Charlson comorbidity index** (median, [IQR]) | 0 [0, 0] | 0 [0, 0] |
| **Liver disease** | 8 (0.4%) | 28 (0.3%) |
| **Kidney disease** | 5 (0.3%) | 16 (0.2%) |
| **Peripheral artery disease** | 11 (0.6%) | 37 (0.4%) |
| **Diabetes mellitus** | 38 (2.0%) | 133 (1.4%) |
| **Stroke** | 12 (0.6%) | 57 (0.6%) |
| **Chronic obstructive pulmonary disease** | 27 (1.4%) | 69 (0.7%) |
| **Hypertension** | 180 (9.4%) | 775 (8.1%) |

<sup>1</sup>International Standard Classification of Education (ISCED) was converted to an education score of 1-4 where 1 indicates ISCED levels 0-2, 2 indicates ISCED level 3, 3 ISCED levels 5-6, and category 4 equals ISCED levels 7-8. Abbreviations: IQR; interquartile range

### Primary outcome

After a median follow-up of 10 years (IQR: 5.9-17.4), 694 CVD events occurred (cHL 170; comparators 524) and 1,183 individuals died (cHL 328; comparators 855). The cumulative risks of CVD were 4.7% (95% CI: 3.6-5.7) for patients with cHL vs. 2.6% (95% CI: 2.3-2.9) for comparators at 5 years, 8.9% (95% CI: 7.2-10.5) vs. 5.5% (95% CI: 4.9-6.0) at 10 years, and 17% (95% CI: 14.1-19.9) vs. 8.2% (95% CI: 7.4-9) at 15 years (*Figure 2 and accompanying table)*.

**Figure 2:**
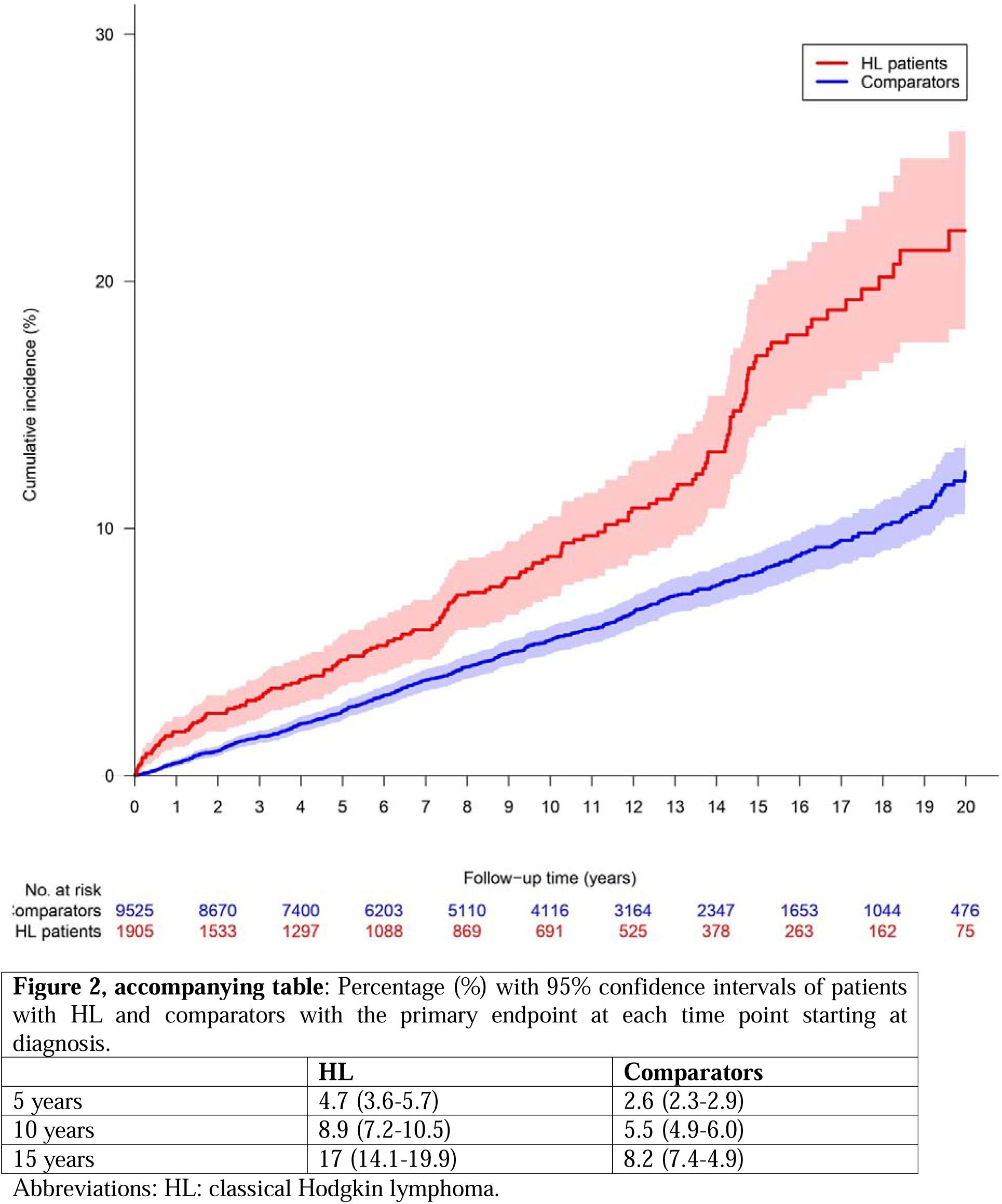
Cause specific cumulative incidence curve and number at risk of the primary endpoint for classical Hodgkin lymphoma patients and matched comparators.

### Secondary outcomes

The most frequent cardiovascular event types for patients with cHL were heart failure, acute coronary syndrome, and atrial arrythmia (*Figure 3 and accompanying table* and *supplementary material S2*). For heart failure, the CIFs were 1.4% (95% CI: 0.8-2) for patients with cHL versus 0.7% (95% CI: 0.6-0.9) for comparators at 5 years, 3.3% (95% CI: 2.2-4.3) vs. 1.5% (95% CI: 1.2-1.8) at 10 years, and 6.5% (95% CI: 4.6-8.4) vs. 2.4% (95% CI: 1.9-2.8) at 15 years.

**Figure 3:**
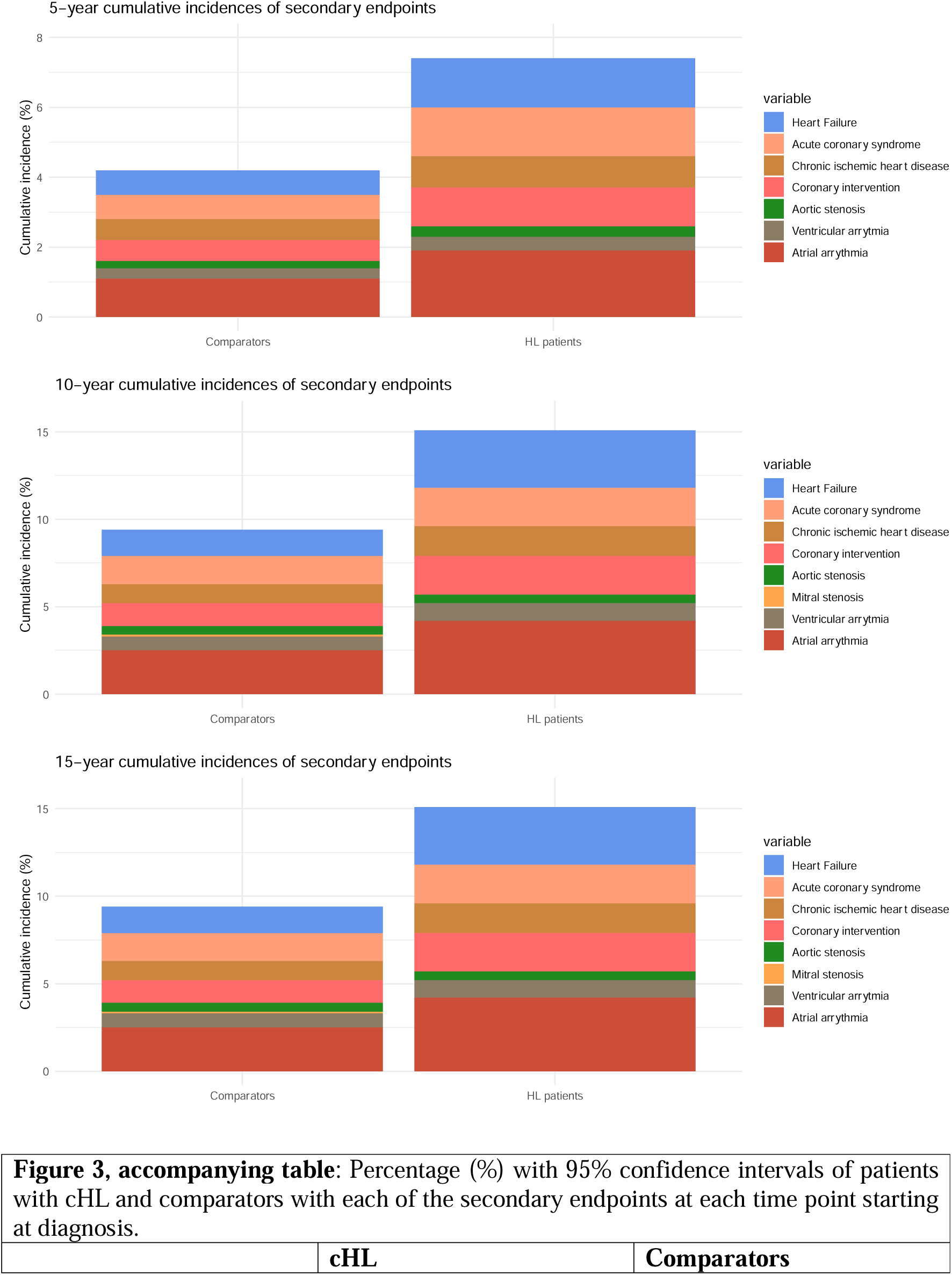
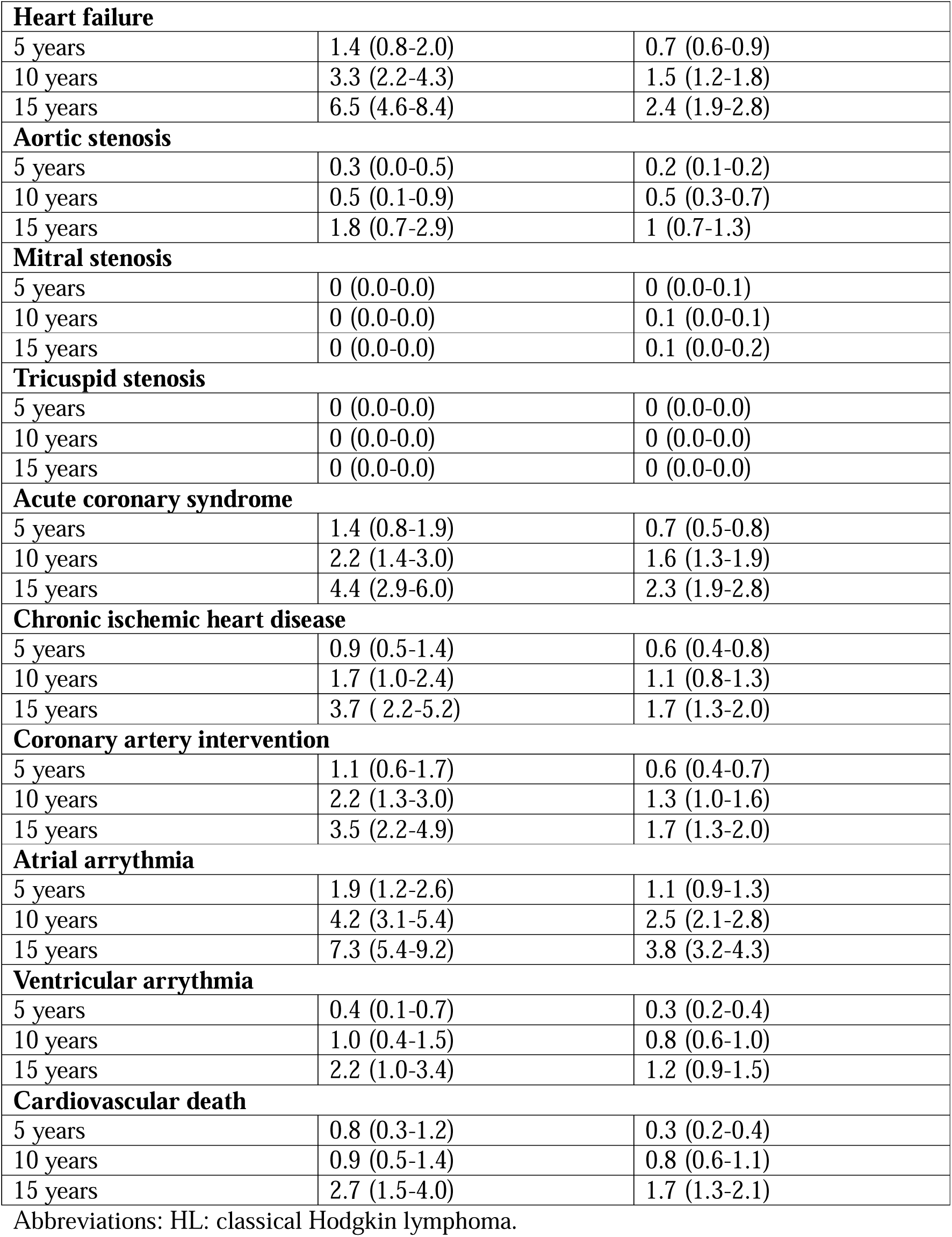
Stacked cumulative incidences at 5-, 10-, and 15-years of the secondary endpoints for Patients with HL and comparators. Please note that each cumulative incidence is based on first event of that diagnosis meaning that an individual can appear in multiple categories.

The risks of aortic valve stenosis were similar between the two groups with 0.3% (95% CI: 0.0-0.5) for patients with cHL vs. 0.2% (95% CI: 0.1-0.2) comparators at 5 years, 0.5% (95% CI: 0.1-0.9) vs. 0.5% (95% CI: 0.3-0.7) at 10 years, and 1.8% (95% CI: 0.7-2.9) vs. 1.0% (95% CI: 0.7-1.3%) at 15 years. Mitral and tricuspid valve stenosis occurred with very low incidences in both groups.

Higher incidences of coronary artery disease were observed among patients with cHL. For acute coronary syndromes the risks were 1.4% (95% CI: 0.9-1.9) for patients with cHL vs. 0.7% (95% CI: 0.5-0.8) for comparators at 5 years, 2.2% (95% CI: 0.8-1.9) vs. 1.6% (95% CI:1.3-1.9) at 10 years, and 4.4% (95% CI: 2.9-6.0) vs. 2.3% (95% CI: 2.8-1.9) at 15 years. For patients with cHL, the risk of coronary intervention was 1.1% (95% CI: 0.6-1.7) at 5 years compared with 0.6% (95% CI: 0.4-0.7) in the comparator group, 2.2% (95% CI:1.3-3.0) vs. 1.3% (95% CI: 0.1-1.6) at 10 years and 3.5% (95% CI: 2.2-4.9) vs 1.7% (95% CI: 1.3-2.0) at 15 years. For chronic ischemic heart disease, the risk was 0.9% (95% CI: 0.5-1.4) vs. 0.6% (95% CI: 0.4-0.8) at 5 years, 1.7% (95% CI: 1.0-2.4) vs. 1.1% (95% CI: 0.8-1.3) at 10 years, and 3.7% (95% CI: 2.2-5.2) vs. 1.7 (95% CI: 1.3-2.0%) at 15 years for patients with cHL and comparators, respectively.

Of the total 1,183 deaths during follow-up, 126 was due to presumed cardiovascular causes (cHL 27; comparators 99). Patients with cHL had a 0.8% (95% CI: 0.3-1.2) risk of death from cardiovascular causes at 5 years compared with 0.3% (95% CI: 0.2-0.4) for comparators, 0.9% (95% CI: 0.5-1.4) vs. 0.8% (95% CI: 0.6-1.1) at 10 years, and 2.7% (95% CI: 1.5-4.0) vs. 1.7% (95% CI: 1.3-2.1) at 15 years.

To examine the impact of surveillance bias and possible diagnosis of pre-existing CVD due to increased diagnostic awareness in the early period after diagnosis of cHL, a sensitivity analysis was performed with start follow-up 90 days after completion of therapy. There was no substantial change in risk estimates for primary endpoints for patients with cHL and comparators when start of follow-up was delayed until 90 days after completion of therapy; 4.0% (95% CI: 3.0-5.0) for patients with cHL vs. 2.9% (95% CI: 2.5-3.2) comparators at 5 years, 8.5% (95% CI: 6.8-10.1) vs. 5.6% (95% CI: 5.1-6.2) at 10 years, and 16.5% (95% CI: 13.6-19.5) vs. 8.6% (95% CI: 7.7-9.4) at 15 years (*Figure 5 and accompanying table*).

### Factors associated with CVD

Hypertension, male sex, and increasing age at diagnosis were associated with development of CVD among patients with cHL (HR for hypertension: 1.82 [95% CI: 1.23-2.71], male sex: 1.64 [95% CI: 1.18-2.27], and age 30-49: 5.57 [95% CI: 2.52-12.32], age 50-69: 18.87 [95% CI: 8.74-40.77], and age >70: 36.41 [95% CI: 15.44-85.88] using age group 18-29 as reference) (*Figure 6*). Cumulative doxorubicin dose was not associated with development of CVD (HR for 200-299 mg/m^2^: 0.85 [95% CI: 0.54-1.32] and HR for ≥300 mg/m^2^: 1.0 [95% CI 0.68-1.48] using 0-199 mg/m^2^ as reference, see also *Figure 4* for association between HR of CVD and cumulative doxorubicin dose). Among comparators, hypertension and male sex had similar increased rates of CVD (HR: 1.84 [95% CI: 1.49-2.27] and 1.93 [95% CI: 1.59-2.34], respectively) (*Supplementary material S3*). Diabetes was not significantly associated with CVD (HR: 1.07, 95% CI: 0.66-1.75) in an analysis adjusted for age, sex, and CCI.

**Figure 4:**
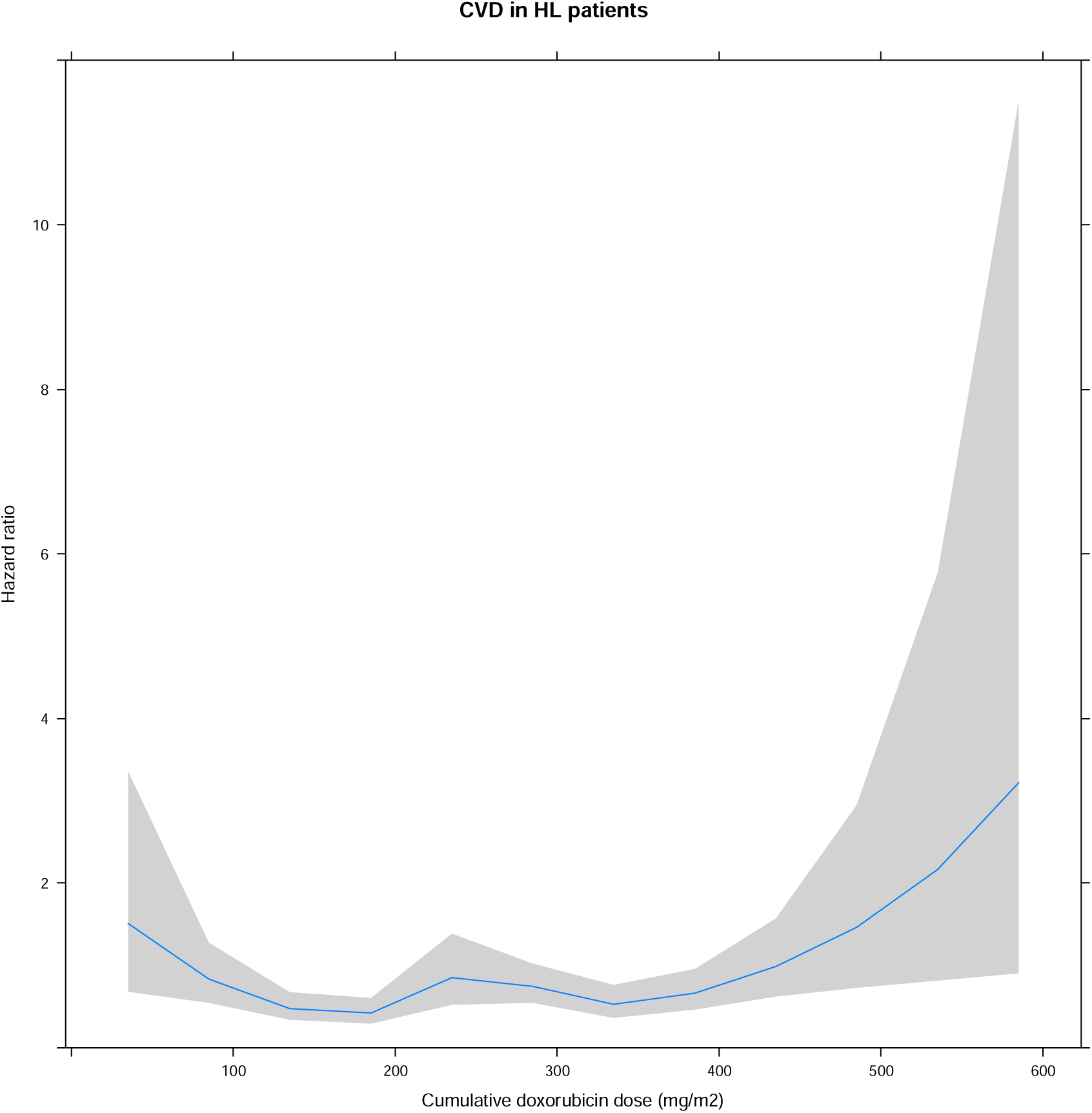
Hazard ratio of the primary endpoint depending on the cumulative doxorubicin dose. Abbreviations: CVD: cardiovascular disease, HL: classical Hodgkin lymphoma.

**Figure 5:**
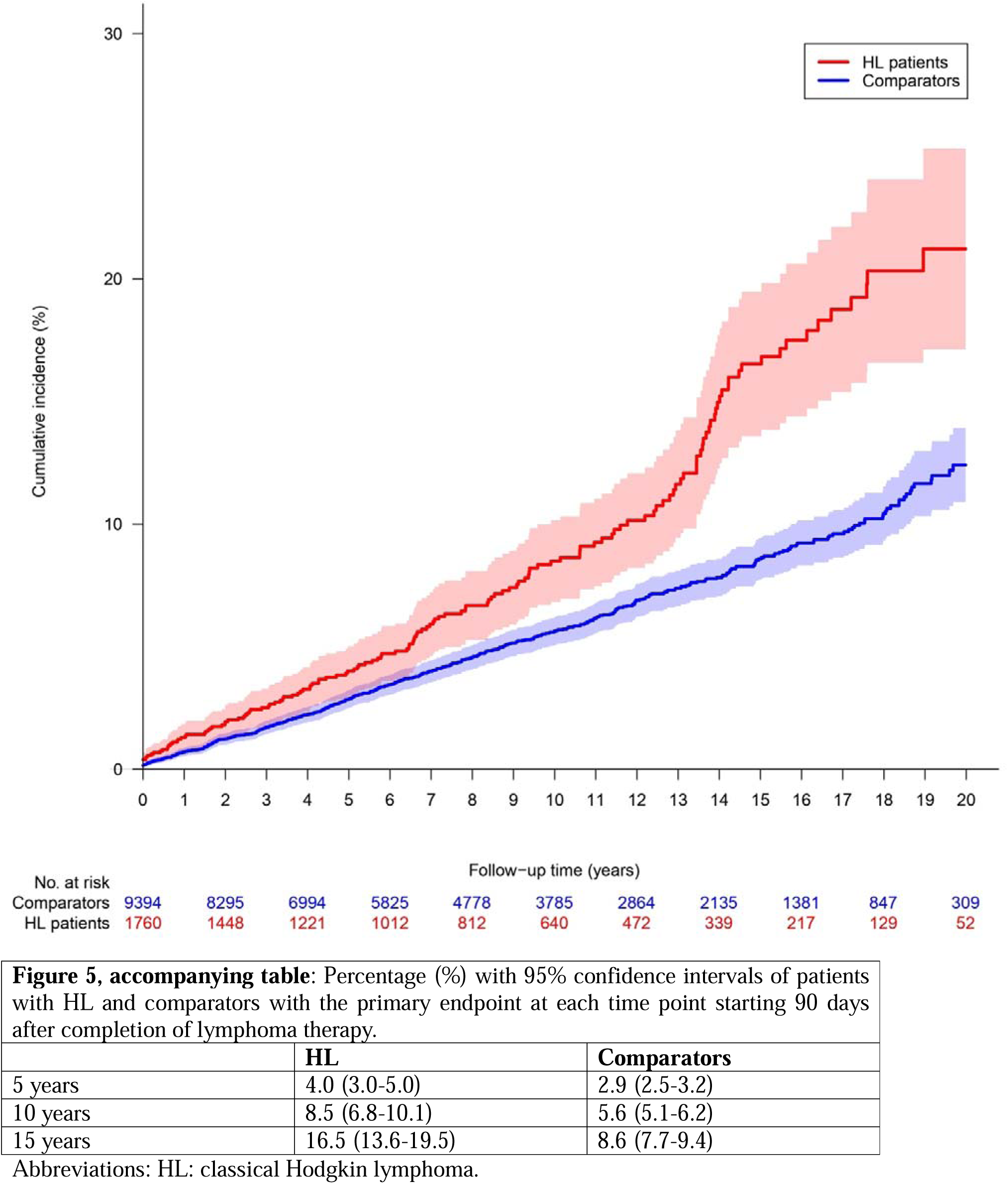
Cause specific cumulative incidence curve and number at risk of the sensitivity analysis with index date (T_0_) set to end of treatment date + 90 days for the primary endpoint for classical Hodgkin lymphoma patients and matched comparators.

**Figure 6:**
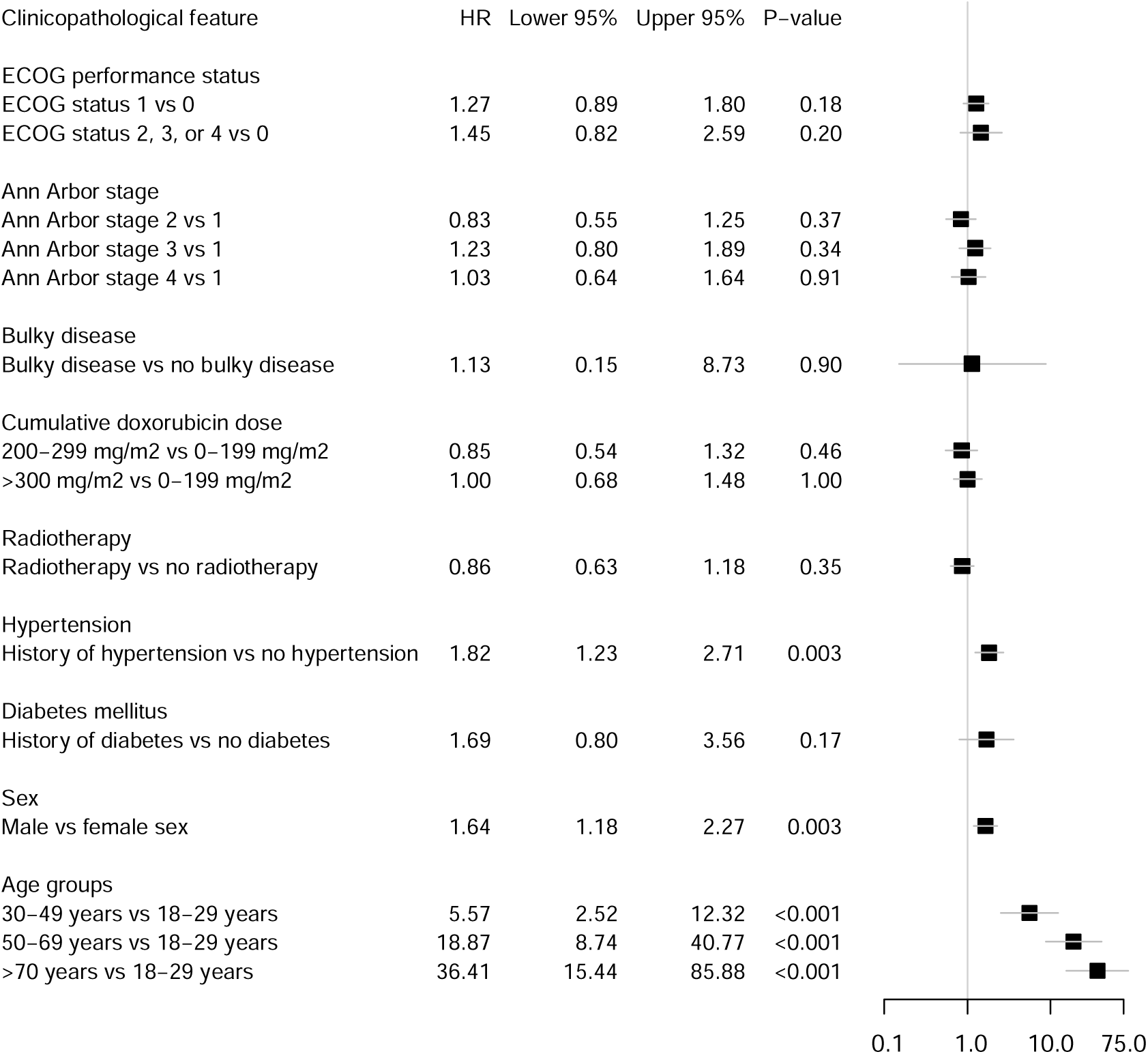
Hazard ratios of the primary outcome in patients with classical Hodgkin lymphoma comparing different clinicopathological features. Abbreviations: HR: hazard ratio, ECOG: Eastern Cooperative Oncology Group performance status

## Discussion

In this nationwide, register-based cohort study on relapse-free patients with cHL treated in first line with ABVD and/or BEACOPP, patients had significantly higher 5-, 10- and 15-year risk of CVD compared with matched comparators from the background population. Well known CVD risk factors such as age, male sex, and hypertension were associated with an increased CVD rate in patients with cHL. Use of radiotherapy was not associated with higher CVD rate among patients, possibly a sign that the improvement in radiotherapy techniques has limited radiation dose to the heart substantially. The main drivers of the CVD risk in patients treated for cHL were heart failure, acute coronary syndromes and atrial arrythmia.

Recent population-based studies of non-Hodgkin lymphoma treated with contemporary treatment regimens have shown substantial risk of heart failure for patients compared to the background population. In a Danish register-based study of patients with aggressive lymphoma treated with high-dose chemotherapy and autologous stem cell transplant, the 10-year cumulative incidence of congestive heart failure was 8.0% versus 2.0% in a matched background population.^29^ A Norwegian study with echocardiographic examination of adult lymphoma survivors treated with autologous hematopoietic stem-cell transplantation found left ventricular dysfunction in 15.7% of patients corresponding to a substantially increased risk compared with controls (odds ratio: 6.6) and a significant proportion of patients were either asymptomatic (5.1%) or only mildly symptomatic (New York Heart Association class II, 8.8%).^30^ Patients with aggressive B-cell lymphomas rarely receive mediastinal radiation with the exception of primary mediastinal B-cell lymphoma prior to the use of PET/CT-adapted consolidation strategies. Therefore, these studies document the cardiotoxic effect of doxorubicin, event when used in doses well below 550 mg/m^2^. Anthracyclines have adverse effects on the myoepithelium, and the type of cardiotoxicity can be classified by time from anthracycline administration until presentation of cardiovascular disease. First, acute toxicity can result in transient arrythmias, non-specific ST-T segment changes on an ECG, or chest pain due to peri-myocarditis and it is estimated to occur in 11% of patients.^31–33^ Rarely, acute toxicity can result in reversible acute left ventricular failure. Second – and most common – is the chronic anthracycline-induced cardiomyopathy which presents within one year after first exposure to antracylines.^34–36^ The last type is the late-onset effects presenting more than one year after first exposure, mainly heart failure or arrythmias.^37–40^ While excess risk identified in this study was modest during the first 10 years, the risk difference after 15 years of follow-up was clinically significant with excess risk of 7.8 percentage points for patients with cHL. A Dutch cohort of 2,524 patients with cHL found a 6.8-fold standardized incidence ratio of developing heart failure compared with expected rates from the general population, corresponding to 58 excess cases per 10,000 person years.^15^ The patients in that study were diagnosed and treated in the years 1965 through 1995 – time periods which were characterized by treatment policies that are very different than today, both in terms of chemotherapy use and radiation techniques. The lower rate of CVD and heart failure in the present study (17% and 6.5% at 15 years, respectively) compared with the Dutch study might reflect these improvements and a successful reduction, although still not complete mitigation, of late CVD toxicities in patients with cHL.

Previously, mediastinal irradiation has consistently been associated with increased risk of coronary artery disease, congestive heart failure and valvular disorders.^3,9,41^ For example, Boivin et al. performed an autopsy-based study of 4,665 patients with cHL diagnosed between 1942-1985 in the United States and reported a 1.87 relative risk of death with any coronary artery disease after mediastinal radiation.^3^ Radiotherapy was not among the factors associated with a higher risk of CVD in Patients with cHL in the present analysis. Unfortunately, location of radiation field is not reported to LYFO, only dose and number of fractions as well as the type of radiotherapy. Previously 35 Gy has been suggested as the lower limit for radiation-induced CVD^42^ and the median Gy applied in this study were 30. Less than 1/3 of patients in the present study received ≥35 Gy and 97% received involved field or involved node radiotherapy. Mantle field was not registered for any patient. Hull et al. reported radiation treatment with a mantle field associated with increased risk of coronary artery disease compared with treatment with a more limited field (HR 7.8, p=0.4).^9^ Both involved field and involved node radiotherapy regimens deliver substantially lower radiation doses to cardiac structures than the traditional mantle field technique^7,43^, possibly explaining the lack of association between radiotherapy and development of CVD. Additive, or possible supra-additive effect of the combination of mediastinal radiation therapy and anthracycline-containing chemotherapy has been suggested.^44^ The treatment algorithms used during the surveyed time were dominated by use of short chemotherapy and radiotherapy for limited stage disease and longer chemotherapy with PET/CT adapted use of radiotherapy for advanced stage disease. Thus, the possible synergistic effects of mediastinal radiotherapy and anthracycline dose will be difficult to explore today due to the often mutually exclusive use of high cumulative doses of anthracyclines and radiotherapy. However, it may to some degree explain the lack of association between radiotherapy use and CVD in the present study.

A prospective clinical study including 339 patients with Hodgkin disease diagnosed between 1964-1992, who were invited for regular visits with special heart exanimations for one year after diagnosis, suggested that the presence of traditional CVD risk factors such as smoking, hypertension, obesity, dyslipidemia, or diabetes among patients with Hodgkin disease led to a significantly higher-than-expected incidence of ischemic heart disease compared with the anticipated risk in the general population with an identical risk profile (p<0.001).^11^ This implies that the presence of CVD risk factors pose a greater risk to patients with cHL than to the general population and that excess risk could be potentially modified by addressing more attention to the modifiable risk factors such as smoking, obesity, hypertension, dyslipidemia, and diabetes mellitus. As expected, and in line with previous studies,^4,9,14^ the present study also found conventional risk factors such as hypertension, age, and male sex significantly associated with increased risk of CVD in patients with cHL. However, for the associations between hypertension and sex and CVD were similar to those observed for comparators suggesting that there is no strong interaction between cHL treatment and those features. Nevertheless, since patients with cHL face an overall higher risk of CVD events, a similar magnitude HR may still cause more events and the impact of well controlled hypertension may be associated with a lower number needed to treat than for comparators. Diabetes was not associated with CVD events for patients with cHL or for comparators. However, given that this study focused on a disease in predominantly younger patients and required at least one ABVD or BEACOPP for study inclusion, the number of elderly patients with diabetes was small (8% of patients ≥70 years) and the power to detect true associations would be limited. Consistent with this, too few patients had hypercholesterolemia to include this well-known risk factor in the analyses. Even though the present study did not document impact of diabetes and hypercholesterolemia on CVD risk in patients with cHL, it is still plausible that patients would benefit substantially from lifestyles that lower risk of these conditions substantially or treatment of those when present according to guidelines on their overall CVD risk.

The main strengths of the present study are the relatively large cohort considering the rarity of the disease, and the maturity of the data ensuring long follow-up for most subjects. LYFO did not contain information of dose of doxorubicin, which means that impact of dose reductions could not be assessed. It is plausible that the dose of doxorubicin was reduced in patients with several risk factors for CVD. However, given the age of the population, the exclusion of patients with CVD events prior to cHL diagnosis, and the restriction to use of at least one ABVD or BEACOPP, it is unlikely that dose reduction was done upfront. Furthermore, the study estimated that BEACOPP contained 35 mg/m^2^ doxorubicin per cycle as in escalated BEACOPP and not standard BEACOPP as this is rarely used. However, this information was not available in LYFO. Lastly, the registries do not contain information on lifestyle factors such as smoking history or body mass index.

Finally, the study is limited by the nature of the data. The registers only include official diagnoses made in a hospital or out-patient clinics and in the previously mentioned Norwegian study, 5.1% of non-Hodgkin lymphoma patients had asymptomatic cardiac damage.^30^ Thus, this study could be at risk of underestimating the incidence of CVD in patients with cHL.

In this nationwide observational cohort study including patients with cHL in long-term remission treated with doxorubicin-containing chemotherapy, cHL patients were still at a substantially higher risk of CVD compared with matched comparators from the background population. This suggests that the use of modern radiotherapy has not eliminated the excess risk of CVD and clinicians should still be aware of the CVD risk in patients with cHL and pay special attention to the modifiable risk factors such as hypertension, dyslipidemia, and diabetes.

## Disclosures

The remaining authors declare no conflicts of interest.

SJG reports grant from the Danish Cancer Association (R327-A18892).

CTP reports grants from Novo Nordisk not related to current study.

HY reports grants from TrygFonden not related to current study.

## Acknowledgments

The authors would like to acknowledge the Region’s Clinical Quality Program (RKKP) for providing access to the Danish Lymphoma Database.

## Data availability statement

The data underlying this article cannot be shared publicly due to ethical and privacy restrictions. The data will be shared on reasonable request to the corresponding author with permission of Statistics Denmark.

**Supplementary material S2**: Cause specific cumulative incidence functions of the secondary endpoints for Hodgkin lymphoma patients and matched comparators.

Abbreviations: HL: classical Hodgkin lymphoma, PCI: percutaneous coronary intervention, CABG: coronary artery bypass grafting, ICD: implantable cardioverter-defibrillator.

**Supplementary material S3:** Hazard ratios of the primary outcome in the comparator group comparing different clinicopathological features.

Abbreviations: HR: hazard ratio.

## Notes

### Competing Interest Statement

The authors have declared no competing interest.

### Funding Statement

This study was funded by the Danish Cancer Association (grant number R327-A18892).

## References

1. Canellos GP, Rosenberg SA, Friedberg JW, et al. Treatment of Hodgkin lymphoma: a 50-year perspective. J Clin Oncol 2014; 32: 163–8.

2. Johnson P, Federico M, Kirkwood A, et al. Adapted Treatment Guided by Interim PET-CT Scan in Advanced Hodgkin’s Lymphoma. New England Journal of Medicine 2016; 374: 2419–2429.

3. Boivin JF, Hutchison GB, Lubin JH, et al. Coronary artery disease mortality in patients treated for Hodgkin’s disease. Cancer 1992; 69: 1241–7.

4. Reinders JG, Heijmen BJ, Olofsen-van Acht MJ, et al. Ischemic heart disease after mantlefield irradiation for Hodgkin’s disease in long-term follow-up. Radiother Oncol 1999; 51: 35–42.

5. Allavena C, Conroy T, Aletti P, et al. Late cardiopulmonary toxicity after treatment for Hodgkin’s disease. Br J Cancer 1992; 65: 908–912.

6. van Nimwegen FA, Ntentas G, Darby SC, et al. Risk of heart failure in survivors of Hodgkin lymphoma: effects of cardiac exposure to radiation and anthracyclines. Blood 2017; 129: 2257–2265.

7. Maraldo M V, Brodin NP, Vogelius IR, et al. Risk of developing cardiovascular disease after involved node radiotherapy versus mantle field for Hodgkin lymphoma. Int J Radiat Oncol Biol Phys 2012; 83: 1232–7.

8. Heidenreich PA, Hancock SL, Lee BK, et al. Asymptomatic cardiac disease following mediastinal irradiation. J Am Coll Cardiol 2003; 42: 743–749.

9. Hull MC. Valvular Dysfunction and Carotid, Subclavian, and Coronary Artery Disease in Survivors of Hodgkin Lymphoma Treated With Radiation Therapy. JAMA 2003; 290: 2831.

10. Adams MJ, Lipsitz SR, Colan SD, et al. Cardiovascular Status in Long-Term Survivors of Hodgkin’s Disease Treated With Chest Radiotherapy. Journal of Clinical Oncology 2004; 22: 3139–3148.

11. Glanzmann C, Huguenin P, Lütolf UM, et al. Cardiac lesions after mediastinal irradiation for Hodgkin’s disease. Radiother Oncol 1994; 30: 43–54.

12. Heidenreich PA, Hancock SL, Vagelos RH, et al. Diastolic dysfunction after mediastinal irradiation. Am Heart J 2005; 150: 977–82.

13. Avilés A, Neri N, Nambo MJ, et al. Late cardiac toxicity secondary to treatment in Hodgkin’s disease. A study comparing doxorubicin, epirubicin and mitoxantrone in combined therapy. Leuk Lymphoma 2005; 46: 1023–1028.

14. Aleman BMP, van den Belt-Dusebout AW, De Bruin ML, et al. Late cardiotoxicity after treatment for Hodgkin lymphoma. Blood 2007; 109: 1878–1886.

15. van Nimwegen FA, Schaapveld M, Janus CPM, et al. Cardiovascular disease after Hodgkin lymphoma treatment: 40-year disease risk. JAMA Intern Med 2015; 175: 1007–17.

16. Jaworski C, Mariani JA, Wheeler G, et al. Cardiac Complications of Thoracic Irradiation. J Am Coll Cardiol 2013; 61: 2319–2328.

17. Specht L. Reappraisal of the role of radiation therapy in lymphoma treatment. Hematol Oncol 2023; 41 Suppl 1: 75–81.

18. Eichenauer DA, Aleman BMP, André M, et al. Hodgkin lymphoma: ESMO Clinical Practice Guidelines for diagnosis, treatment and follow-up. Annals of Oncology 2018; 29: iv19–iv29.

19. Lefrak EA, Pitha J, Rosenheim S, et al. A clinicopathologic analysis of adriamycin cardiotoxicity. Cancer 1973; 32: 302–14.

20. Baech J, Hansen SM, Lund PE, et al. Cumulative anthracycline exposure and risk of cardiotoxicity; a Danish nationwide cohort study of 2440 lymphoma patients treated with or without anthracyclines. Br J Haematol 2018; 183: 717–726.

21. Schmidt M, Pedersen L, Sørensen HT. The Danish Civil Registration System as a tool in epidemiology. Eur J Epidemiol 2014; 29: 541–9.

22. Pedersen CB. The Danish Civil Registration System. Scand J Public Health 2011; 39: 22–25.

23. Arboe B, El-Galaly TC, Clausen MR, et al. The Danish National Lymphoma Registry: Coverage and Data Quality. PLoS One 2016; 11: e0157999.

24. Lynge E, Sandegaard JL, Rebolj M. The Danish National Patient Register. Scand J Public Health 2011; 39: 30–33.

25. Wallach Kildemoes H, Toft Sørensen H, Hallas J. The Danish National Prescription Registry. Scand J Public Health 2011; 39: 38–41.

26. Helweg-Larsen K. The Danish Register of Causes of Death. Scand J Public Health 2011; 39: 26–29.

27. Jensen VM, Rasmussen AW. Danish Education Registers. Scand J Public Health 2011; 39: 91–4.

28. Andersen MP, Valeri L, Starkopf L, et al. The Mediating Effect of Pupils’ Physical Fitness on the Relationship Between Family Socioeconomic Status and Academic Achievement in a Danish School Cohort. Sports Medicine 2019; 49: 1291–1301.

29. Baech J, Husby S, Trab T, et al. Cardiovascular diseases after high-dose chemotherapy and autologous stem cell transplant for lymphoma: A Danish population-based study. Br J Haematol 2024; 204: 967–975.

30. Murbraech K, Smeland KB, Holte H, et al. Heart Failure and Asymptomatic Left Ventricular Systolic Dysfunction in Lymphoma Survivors Treated With Autologous Stem-Cell Transplantation: A National Cross-Sectional Study. Journal of Clinical Oncology 2015; 33: 2683–2691.

31. Ferrans VJ. Overview of cardiac pathology in relation to anthracycline cardiotoxicity. Cancer Treat Rep 1978; 62: 955–61.

32. Steinberg JS, Cohen AJ, Wasserman AG, et al. Acute arrhythmogenicity of doxorubicin administration. Cancer 1987; 60: 1213–8.

33. Chatterjee K, Zhang J, Honbo N, et al. Doxorubicin cardiomyopathy. Cardiology 2010; 115: 155–62.

34. Friedman MA. Doxorubicin cardiotoxicity. Serial endomyocardial biopsies and systolic time intervals. JAMA: The Journal of the American Medical Association 1978; 240: 1603–1606.

35. Bristow MR, Billingham ME, Mason JW, et al. Clinical spectrum of anthracycline antibiotic cardiotoxicity. Cancer Treat Rep 1978; 62: 873–9.

36. Von Hoff DD, Rozencweig M, Layard M, et al. Daunomycin-induced cardiotoxicity in children and adults. A review of 110 cases. Am J Med 1977; 62: 200–8.

37. Larsen RL, Jakacki RI, Vetter VL, et al. Electrocardiographic changes and arrhythmias after cancer therapy in children and young adults. Am J Cardiol 1992; 70: 73–7.

38. Yeung ST, Yoong C, Smith PJ, et al. Functional myocardial impairment in children treated with anthracyclines for cancer. The Lancet 1991; 337: 816–818.

39. Lipshultz SE, Colan SD, Gelber RD, et al. Late Cardiac Effects of Doxorubicin Therapy for Acute Lymphoblastic Leukemia in Childhood. New England Journal of Medicine 1991; 324: 808–815.

40. Steinherz LJ, Steinherz PG, Tan CT, et al. Cardiac toxicity 4 to 20 years after completing anthracycline therapy. JAMA 1991; 266: 1672–7.

41. Hancock SL, Tucker MA, Hoppe RT. Factors affecting late mortality from heart disease after treatment of Hodgkin’s disease. JAMA 1993; 270: 1949–55.

42. Ng AK. Late complications after treatment for Hodgkin lymphoma. Curr Hematol Malig Rep 2008; 3: 119–125.

43. Koh E-S, Tran TH, Heydarian M, et al. A comparison of mantle versus involved-field radiotherapy for Hodgkin’s lymphoma: reduction in normal tissue dose and second cancer risk. Radiation Oncology 2007; 2: 13.

44. Myrehaug S, Pintilie M, Tsang R, et al. Cardiac morbidity following modern treatment for Hodgkin lymphoma: Supra-additive cardiotoxicity of doxorubicin and radiation therapy. Leuk Lymphoma 2008; 49: 1486–1493.

