## Supplementary material S1 for "Risk of cardiovascular disease in patients with classical Hodgkin lymphoma: a Danish nationwide register-based cohort study"

### Disease definitions

| **Variable** | **Identifier** |
| --- | --- |
| Primary outcome, composite of cardiovascular diseases | Constrictive pericarditis: ICD-10: DI310, DI311 |
|  | Aortic stenosis:  ICD-10: DI350, DI352 NOMESCO: KFMA, KFMD |
|  | Mitral stenosis:  ICD-10: DI342  NOMESCO: KFKA, KFKD |
|  | Tricuspid stenosis:  ICD-10: DI360, DI362  NOMESCO: KFGA, KFGE |
|  | Heart failure:  ICD-10: DI50*, DI110, DI130, DI132, DZ950B, DI421, DI425, DI420, DI428, DI429 |
|  | Ischemic heart disease:  ICD-10: DI208, DI209, DI249, DI251, DI255, DI258, DI250 |
|  | Acute myocardial infarction:  ICD-10: DI21*, DI200 |
|  | Coronary interventions:  NOMESCO: KFNG, KFNF, KFNA, KFNB, KFNC, KFND, KFNE |
|  | Atrial arrythmias: DI48* |
|  | Ventricular arrythmias:  ICD-10: DI490, DI46* DI472, DZ950D  NOMESCO: BFCB0, BFCB6 |
| Constrictive pericarditis | ICD-10: DI310, DI311 |
| Aortic valve stenosis | ICD-10: DI350, DI352 NOMESCO: KFMA, KFMD |
| Mitral valve stenosis | ICD-10: DI342  NOMESCO: KFKA, KFKD |
| Tricuspid valve stenosis | ICD-10: DI360, DI362  NOMESCO: KFGA, KFGE |
| Heart failure | ICD-10: DI50*, DI110, DI130, DI132, DZ950B, DI420, DI428, DI429, DI421, DI425 |
| Chronic ischemic heart disease | ICD-10: DI208, DI209, DI249, DI251, DI255, DI258, DI250 |
| Acute coronary syndromes | ICD-10: DI21*, DI200 |
| Coronary interventions | NOMESCO: KFNG, KFNF, KFNA, KFNB, KFNC, KFND, KFNE |
| Atrial arrythmia | ICD-10: DI48* |
| Ventricular arrythmia | ICD-10: DI490, DI46* DI472, DZ950D  NOMESCO: BFCB0, BFCB6 |

Abbreviations: ICD-10: International Classification of Diseases system 10^th^ revision, NOMESCO: Nordic Medico Statistical Committee classification
