## Supplementary figures and images for "Risk of cardiovascular disease in patients with classical Hodgkin lymphoma: a Danish nationwide register-based cohort study"

### Supplementary material S2

#### Supplementary material S2

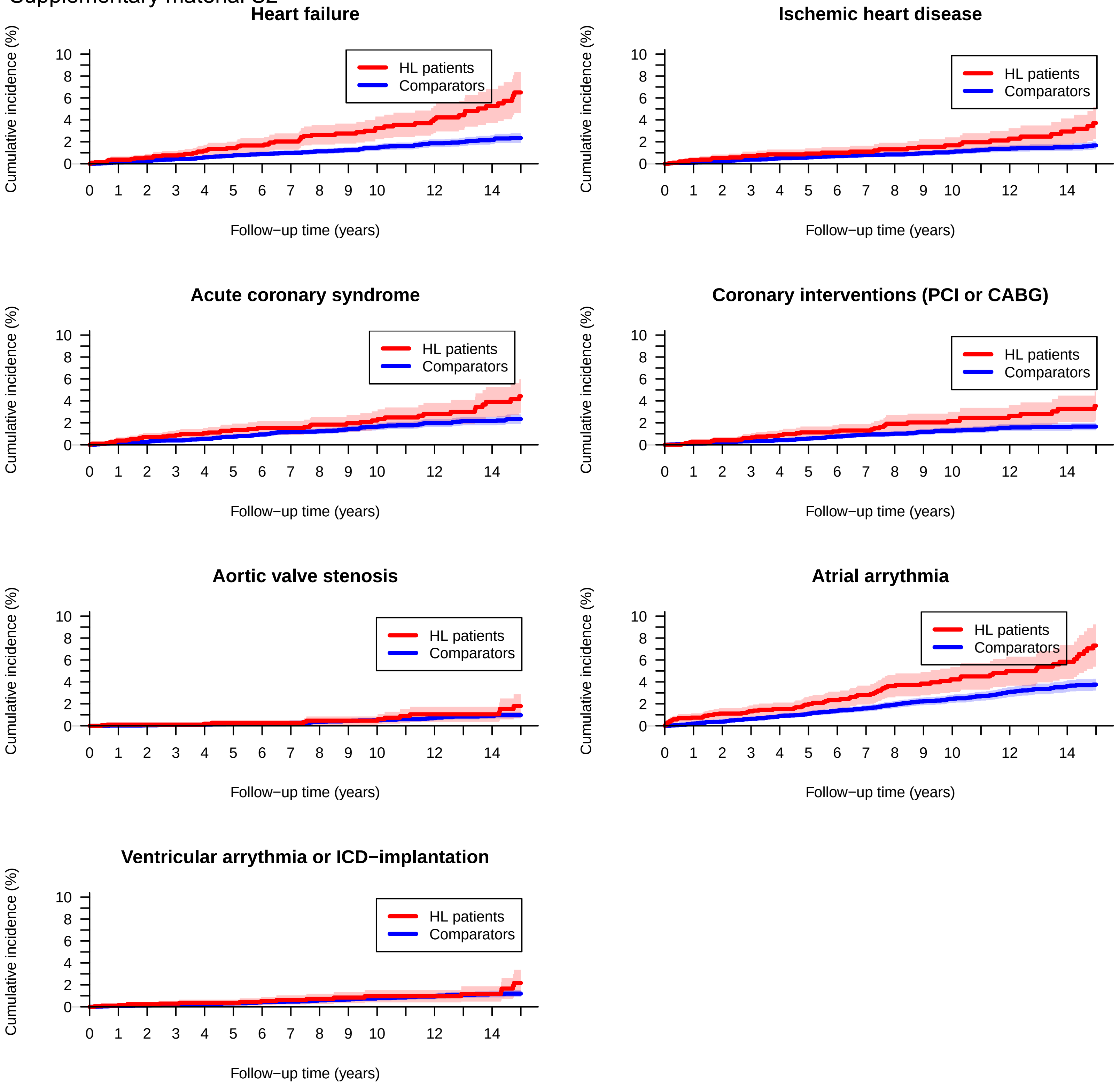
