## Supplementary material S3 for "Risk of cardiovascular disease in patients with classical Hodgkin lymphoma: a Danish nationwide register-based cohort study"

| Clinicopathological feature | HR | Lower 95% | Upper 95% | P−value |
|---|---|---|---|---|
| Hypertension | | | | |
| History of hypertension vs no hypertension | 1.84 | 1.49 | 2.27 | 0.003 |
| Diabetes mellitus | | | | |
| History of diabetes vs no diabetes | 1.07 | 0.66 | 1.75 | 0.78 |
| Sex | | | | |
| Male vs female sex | 1.93 | 1.59 | 2.34 | 0.003 |
| Age groups | | | | |
| 30−49 years vs 18−29 years | 2.88 | 1.72 | 4.81 | <0.001 |
| 50−69 years vs 18−29 years | 14.30 | 8.86 | 23.09 | <0.001 |
| >70 years vs 18−29 years | 33.74 | 20.37 | 55.88 | <0.001 |

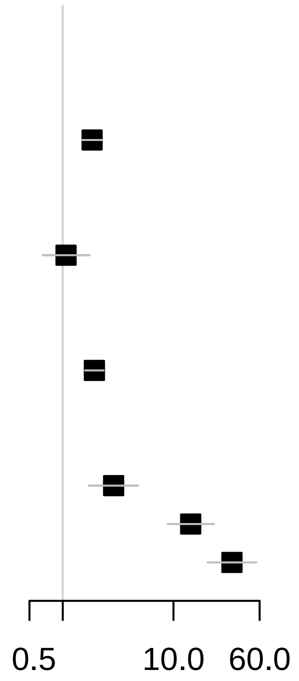
